# The clots removed from ischaemic stroke patients by mechanical thrombectomy are amyloid in nature

**DOI:** 10.1101/2024.11.01.24316555

**Authors:** Justine M. Grixti, Arun Chandran, Jan-Hendrik Pretorius, Melanie Walker, Alakendu Sekhar, Etheresia Pretorius, Douglas B Kell

## Abstract

Ischemic stroke due to large vessel occlusion results from the blockage of a major cerebral artery by a clot; however, the origins and molecular composition of these clots remain poorly understood. Mechanical thrombectomy has become a standard treatment to remove obstructive clots, providing a unique opportunity to analyze their properties. We previously demonstrated that blood can clot into an amyloid-like form, generating fibrinaloid microclots (2–200 μm) that are highly resistant to fibrinolysis. In this study, archived clots from eight ischemic stroke patients with large vessel occlusion were examined, using samples stored in the Walton Centre Clot Bank in Liverpool, UK. All clots exhibited strong, heterogeneous amyloid staining, revealing a pervasive amyloid component. These findings represent a previously unreported characteristic of stroke clots, highlighting the potential for amyloid-targeted therapies to overcome their fibrinolytic resistance and providing a foundational new insight into ischaemic stroke pathophysiology and treatment.

## Introduction

Stroke is the second leading cause of death worldwide, with an annual mortality rate exceeding 5 million persons (Donkor 2018; GBD 2019 Stroke Collaborators 2021; He et al. 2024; Powers 2020). In the more common ischaemic stroke, a blood vessel that supplies the brain becomes blocked by a clot or thrombus of some kind. A common and effective treatment is early enzymatic thrombolysis (Al-Ajlan et al. 2024; Gurman et al. 2015; Wardlaw et al. 2014), but this is not always possible and is not without a risk of inducing bleeding due to a lack of specificity of the relevant enzymes for fibrin (Emberson et al. 2014; Gurman et al. 2015; Röther et al. 2013; Zhong et al. 2024; Zubair and Sheth 2023). Endovascular mechanical thrombectomy to remove the blood clot responsible for the stroke is also now widespread (Campbell et al. 2019; El Tawil and Muir 2017; Hurford et al. 2020; Martin-Gutiérrez et al. 2024; Menon et al. 2019; Powers 2020), alone or additionally (Hammed et al. 2024; Heiferman et al. 2017; Rossi et al. 2022; Suzuki et al. 2021), and has the considerable scientific advantage that one can then study the clots so extracted (Huang and Bhaskar 2022; Martha et al. 2023; Walker et al. 2022). Some time ago, we discovered (Kell and Pretorius 2017; Pretorius et al. 2016) that blood can clot into an anomalous ‘fibrin amyloid’ or ‘fibrinaloid’ (Kell et al. 2022; Kell and Pretorius 2023; Nunes et al. 2022; Turner et al. 2023a) form that is relatively resistant to fibrinolysis. This would of course explain why such clots are not removed naturally (Desilles et al. 2022; Ho-Tin-Noé et al. 2023), but the clots there observed were microclots, whose equivalent diameter is commonly in the range 2-200 mm (Bergaglio et al. 2024; Dalton et al. 2024; Grobbelaar et al. 2022; Grobbelaar et al. 2021; Kell et al. 2022; Kruger et al. 2022; Nunes et al. 2022; Pretorius and Kell 2024; Pretorius et al. 2022; Schofield et al. 2024; Turner et al. 2023b). The availability of clots removed by thrombectomy here allows us to assess whether the mm-sized clots obtained following ischaemic stroke are ‘normal’, or whether they are in fact amyloid in character. We show that they are highly amyloid in nature.

We here assess the amyloid nature of the thrombectomised samples using standard fluorogenic staining. Of the many amyloid stains available (Chisholm and Hunter 2024), the most commonly used is thioflavin T, since its fluorescence is quenched (via an intramolecular bond rotation) (Amdursky et al. 2012) when in solution, but it is highly fluorescent when bound to an amyloid target (Biancalana and Koide 2010). Although a variety of other stains have been used to assess the clot morphology (and indeed origin) (Huang et al. 2022; Huang and Bhaskar 2022), we know of no study that has sought to stain for amyloid. This is what we do here, with striking results.

## Methods

### Thrombectomy, patient consenting, ethics and wax embedding

Thrombectomies were performed using flow arrest with aspiration, and/or using the stent retriever technique, under general anaesthesia. The Walton center thrombectomy clot bank pilot feasibility trial formalized the clot collection and analysis process for patients with large vessel occlusion (LVO) stroke who had undergone thrombectomy.

The patient cohort enrolled in the clot bank feasibility trial were anonymously assigned a participant number (as in Table 1) and a proforma. Consent was obtained from next of kin (NOK) prior to thrombectomy completion.

**Table 1:**
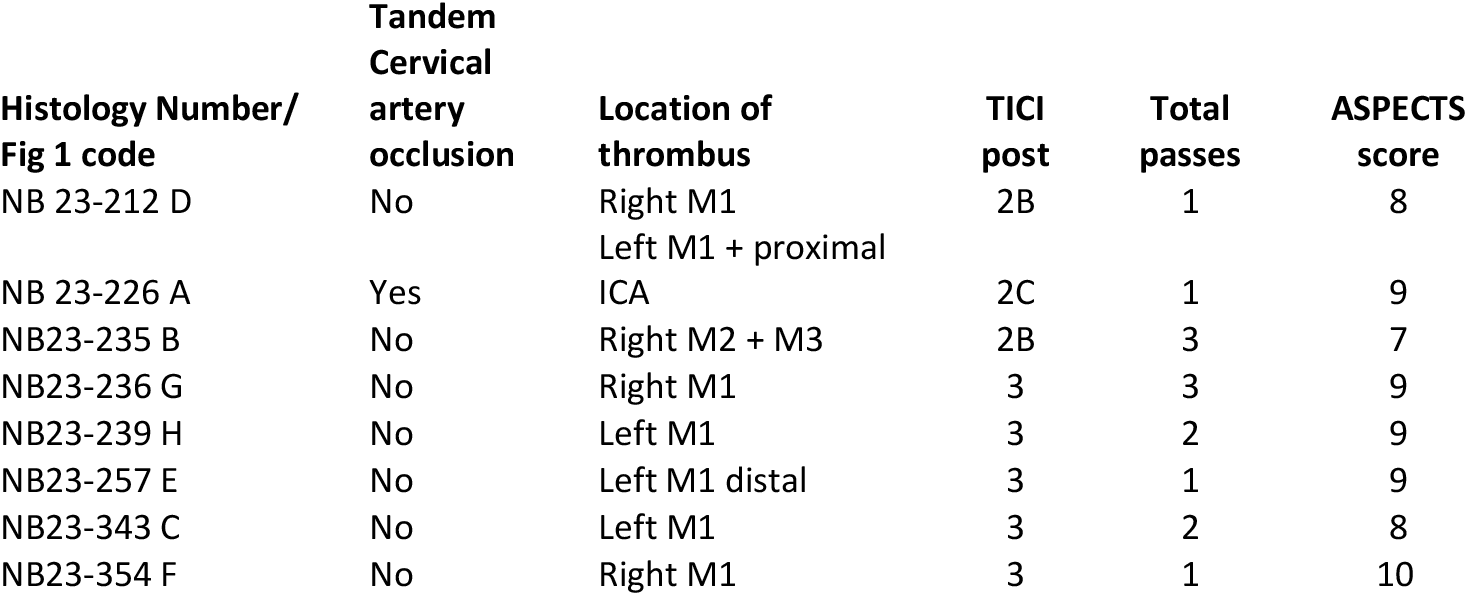
Procedural Characteristics and Outcomes of Retrieved Thrombi. R elevant metadata for the wax-embedded clots extracted from the eight patients studied. The Thrombosis in Cerebral Infarction (TICI) scale (Higashida et al. 2003) is a tool used to grade the degree of perfusion obtained following recanalization of an arterial occlusion (0 meaning no perfusion). The ASPECTS (Alberta Stroke Program Early CT Score) (Mokin et al. 2017) is a quantitative score that measures the extent of early ischemic changes in anterior circulation hyperacute ischemic stroke.

For this specific analysis of clot samples obtained during the clotbank feasibility trial (IRAS 308223) a substantial amendment to cover the present studies was submitted to the Health research authority (HRA) for review by the Research Ethics Committee. A favourable opinion and approval for the amendment was obtained (REC reference 22/NW/0087) before the commencement of the formal analysis of the samples.

#### Study Specimen Collection

During the procedure, the neuro-interventionalist collected the samples, which are removed during the thrombectomy procedure (samples collected between Monday-Thursday 9am-5pm). The retrieved thrombus materials (which would normally be discarded) were collected and stored in sample pots containing 10% formalin and transferred by a member of the neuroradiology team in a manner that did not delay or slow the thrombectomy procedure. The specimen pots were labelled according to the origin of the retrieved clot samples as proximal, distal, specific vessel information, 1st pass, 2nd pass and so on. The labelled formalin pots with patient addressographs were transferred to the Neuropathology lab and incorporated into the existing sample tracking system. Once the patient/personal consultee has consented /agreed to the retrieved clot being kept for future analysis, it was stored in the lab, registered as an ‘LNBW’ sample, and a unique study number generated. 8 out of the 31 patients who were asked (from nine different hospitals) consented during the recruitment period (7 March to 30 April, 2023), and these represent the study sample.

Within this sample, ages ranged from 58 to 88 years old, with all 8 patients being white Caucasian and all but one were males. None of the participants had a recent history of malignancies or infective endocarditis. Medical comorbidities ranged from hypertension, hyperlipidemia, gastric reflux, CABG, Atrial Fibrillation, diabetes, and smoking. All patients accepted by the center for thrombectomy had an mRS (modified Rankin scale) score (Wilson et al. 2005) between 0 and 2 (not shown). The majority of thrombi were found in the middle cerebral artery, with 100% of cases being treated using the flow arrest, aspiration, and stent technique. The longest time from onset of stroke to vessel recanalization seen in our cohort was 10 hours 32 minutes, which was seen in a patient where the stroke occurred overnight. In terms of IV thrombolysis, 80% (n = 6/8; all bar 23-212 and 23-254) received this treatment prior to thrombectomy; clearly it had little obvious effect on the overall clot properties. A specific separate proforma for recording the laboratory data and clot characteristics was used.

### Initial macroscopic examination

Following 24hr formalin fixation, the specimen was examined macroscopically by the pathologists to include descriptors as outlined in the LNBW brain clot bank pathology report (not shown). A macrophotograph of each formalin fixed sample was obtained and stored in the archives.

### Processing, initial staining and microscopic examination at the Walton Centre

A single, tubular fragment was embedded along the length of the thrombus. In case of fragmented specimens all the fragments were embedded in one block. The aim was to embed material so that a maximum cross-section is achieved when sectioning. Paraffin blocks were sectioned at a 3-5μm thickness. Paraffin blocks were trimmed until a full-face section of the clot is reached. Glass slides were labelled with the histology number. Although not reported here, all three levels of tissue sections were stained with Hematoxylin & Eosin (H&E), to assess the overall structure of the clot, and Martius Scarlet Blue (MSB) stains; the latter stain helps to differentiate fibrin from red blood cells (RBCs). Van Gieson staining was performed when required for assessment of collagen content.

ASPECTS were applied to capture the amount of the brain affected during the insult.

### Sectioning, dewaxing and fluorescence microscopy at the University of Liverpool

Microtome sectioning and dewaxing was performed by cooling the clots’ wax blocks on ice, followed by cutting 5 μm sections on a Leica RM2235 microtome. Sections were then de-waxed in xylene for 5 minutes, then hydrated in 100%, 90% and 70% ethanol, respectively. Each section was then transferred to individual wells in a 15-well glass bottom angiogenesis Ibidi slide (Grixti et al. 2024). Fluorescence microscopy using a Cytation 1 instrument and the fluorescent dye Thioflavin T was performed as described in (Grixti et al. 2024), following protocols developed by Dalton and colleagues (Dalton et al. 2024).

## Results

Clots were obtained from the clot bank held in Liverpool, UK, dewaxed as in Methods, exposed to thioflavin T (30 mM), and imaged in a Cytation-1 fluorescence microscope (Grixti et al. 2024). Figure 1 A-H shows slices from typical thrombectomies taken from each of the eight patients/ clots studied. From these striking pictures, it is evident that (i) there is in each case a massive amount of amyloid, shown by the green fluorescence of added thioflavin T, (ii) there is considerable heterogeneity in staining intensity within the clots, and (iii) there is considerable heterogeneity in staining between the clots. Adjacent to each of the micrographs is a histogram plot created using the histogram function in Image J. These show the distribution of pixel intensities for those samples. Again both these histograms and the images themselves show the very extensive heterogeneity of the clots as regards amyloid formation.

**Figure 1:**
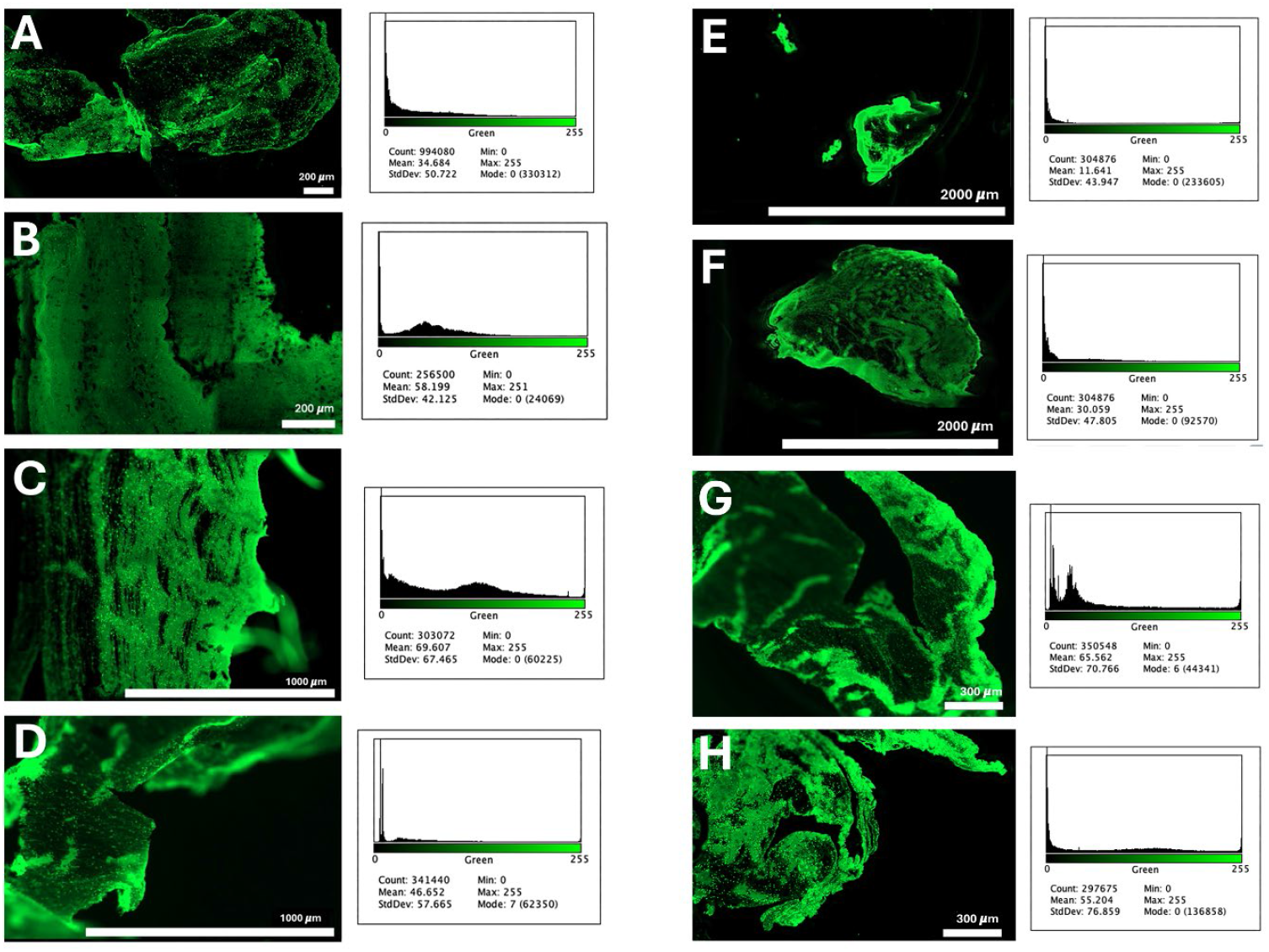
Fluorescence images of 8 thrombectomies representing 8 samples from stroke patients. Samples were embedded in wax followed by sectioning, dewaxing and Thioflavin staining. Adjacent to each of the micrographs is a histogram plot showing the distribution of pixel intensities for each sample. The patients represented as A to H are given in Table 1.

The substantial heterogeneity both within and between such clots has been observed by others (Dumitriu LaGrange et al. 2022; Liu et al. 2022). To quantify this more clearly, we show all the fluorescence intensity distributions of the eight images as distribution plots in Figure 2.

**Figure 2.**
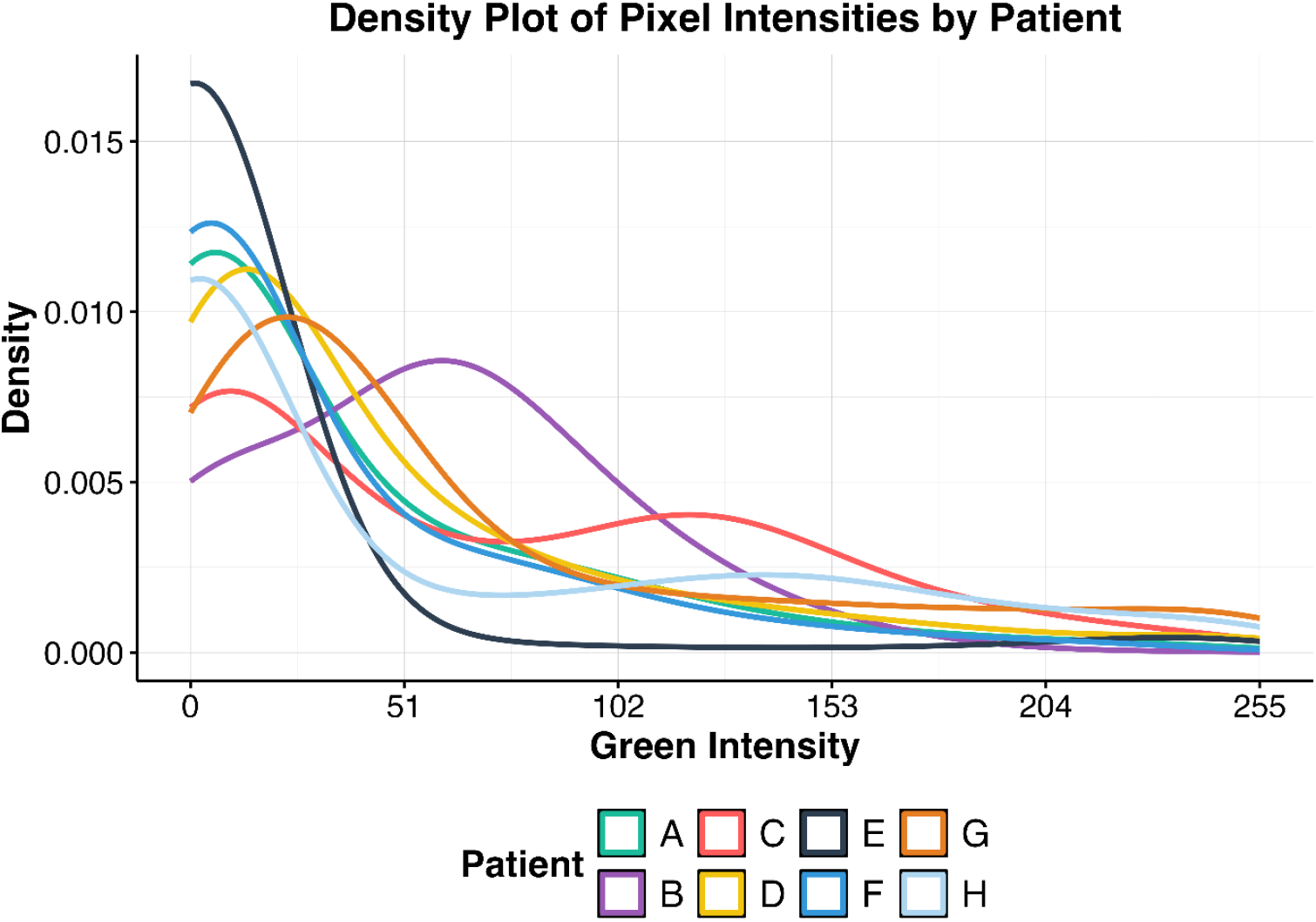
Intensity distributions of the pixel values in the eight images given in Figure 1, illustrating the heterogeneity of staining both within and between samples.

The chief point is that <u>in every single case</u>, regardless of the thrombus location or of other metadata given in Table 1, the extent of amyloid staining is truly massive, with in most cases the majority of the clot being amyloid in character. However, because of the relatively small numbers of clots studied, we do not perform further statistical analyses, and confine ourselves here to the headline message.

While our chief interest here is the very extensive presence of amyloid in these microclots, as judged by the thioflavin T staining, it is of interest to get a feel for the extent of the amyloid distribution in different clots. To this end, using different slices, we also show in Figure 3 two paired brightfield and fluorescence images to illustrate the kind of heterogeneity observed. In one (patient 23-236; A, B) the amyloid is preferentially at the edges of the clot (plausibly explaining its resistance to fibrinolysis), while in the second example (patient 23-354; C, D) the amyloid is present more or less throughout the clot.

**Figure 3:**
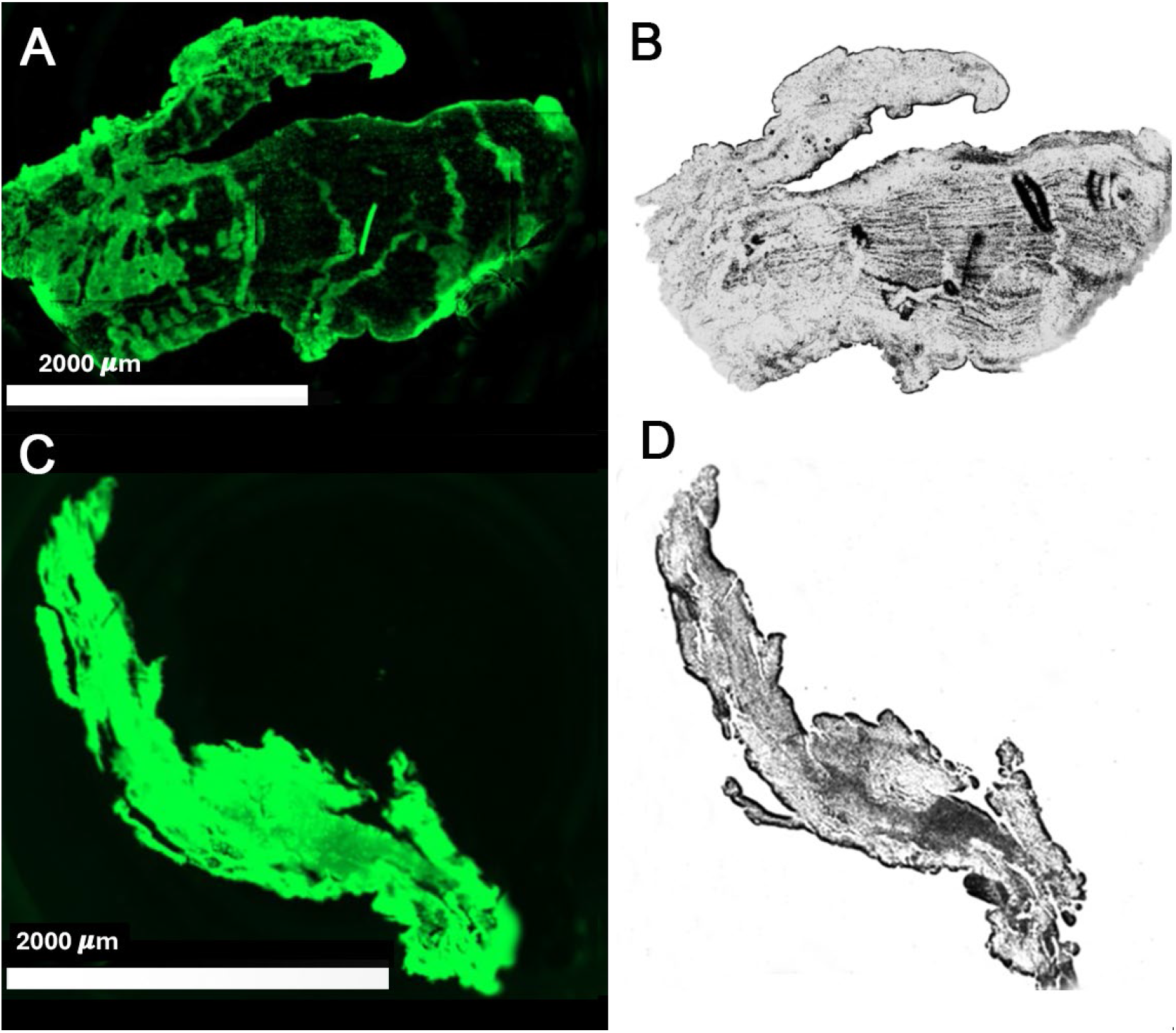
Paired amyloid-stained (A, C) and brightfield (B, D) images of two typical thrombectomised clots, as observed using the Cytation 1. Panels A and B are from Patient 23-236 (numerology as Table 1) while C and D are from patient 23-354.

Wax embedding is a very well-established medium for the detection of ‘classical’ amyloids using birefringent or fluorescent stains (Bély and Makovitzky 2006; Kung et al. 2002; Murphy et al. 2001; Shtrasburg et al. 1997; van de Kaa et al. 1986). Consequently, it is very implausible that these findings might simply be due to the wax embedding.

## Discussion

Most analyses of retrieved thrombi have focused on morphology, specifically classifying thrombi as fibrin-rich or erythrocyte-rich to assist in imaging, procedural planning, and outcome prediction. Recently, studies have expanded to examine additional compositional elements, such as microbial signatures and inflammatory components, that may further influence thrombus morphology. It was observed more than ten years ago using the electron microscope that the clots observable following ischaemic stroke had an anomalous character, that was both denser and more matted than that of normal clots (Pretorius et al. 2011a; Pretorius et al. 2011b; Pretorius et al. 2010). It was subsequently recognised that such ‘dense matted deposits’ (Pretorius et al. 2017a; Pretorius et al. 2013; Swanepoel et al. 2016) are in fact amyloid in nature (Pretorius et al. 2016; Pretorius et al. 2017b). What is clear from the present findings is that the clots extracted from stroke patients by mechanical thrombectomy are themselves considerably, and in some cases almost almost entirely, amyloid in character. This has considerable ramifications in terms of both the scientific understanding of their genesis and possible therapies. Molecules known to induce amyloid-type clotting include bacterial cell wall components (Pretorius et al. 2016; Pretorius et al. 2017b) and the SARS-CoV-2 spike protein (Grobbelaar et al. 2021), which is itself highly amyloidogenic (Larsson et al. 2023; Nyström and Hammarström 2022), and associated with stroke risk (Ab Rahman et al. 2024). It is thus pertinent that both kinds of infection are associated with an increased stroke risk (Westendorp et al. 2011; Zuin et al. 2023). Sepsis also increases both fibrinaloid formation (Schofield et al. 2024) and classical amyloidoses (Basak et al. 2021).

As well as the association of stroke with bacterial cell wall components, it is also of some considerable interest that clots may also in fact contain bacteria (Patrakka et al. 2021; Patrakka et al. 2019; Walker et al. 2022), since we have previously shown that even when present in minuscule concentrations bacterial cell wall products can catalyse the formation of fibrinaloid microclots (Pretorius et al. 2016; Pretorius et al. 2018b), and that oral bacteria are also associated with stoke (Aliena-Valero et al. 2021; Freiherr Von Seckendorff et al. 2024; Leonov et al. 2024; Staessens and De Meyer 2021; Yadav et al. 2023; Zhang et al. 2024a).

It is usually assumed that the clots underpinning the blockages in ischaemic stroke arrive at the sites where they act more or less fully formed (cerebral embolism) or by growing there ‘in one event’ (thrombosis). However, it seems implausible that these clots could all flip from a normal to an amyloid conformation after arriving at such sites, so one has to assume that – like other ‘classical’ amyloid fibrils – they formed by accretion, most plausibly for their amyloid portions from the pre-existing fibrinaloid microclots that have been widely observed in a variety of inflammatory diseases (e.g. (Dalton et al. 2024; Pretorius et al. 2018a; Pretorius and Kell 2024; Pretorius et al. 2017a; Pretorius et al. 2018c)), including Long COVID (Kell et al. 2022; Kell and Pretorius 2023, 2022; Pretorius et al. 2022; Turner et al. 2023b).

Although it is common to give recombinant tPA to stroke patients (Barreto 2011; Emberson et al. 2014) (6 of 8 in the present study received this treatment), it is evident visually (Figure 1) that this must have induced comparatively little macroscopic clot breakdown; those who did not receive tPA (Panels D and F in Fig 1) are not noticeably different in terms of amyloid staining from those that did (Figure 2). Given that tPA is the normal means of initiating fibrinolysis *in vivo* (before the stroke suffered by these individuals), and had evidently failed to break down the amyloid clots, it is possibly unsurprising that adding more had comparatively little effect.

The clots removed from ischaemic stroke patients by thrombectomy vary in size, and their lengths and diameters are correlated (Rossi et al. 2021), which is also consistent with the idea that they do grow by accretion. In that study (Rossi et al. 2021), typical clot lengths depended on the artery involved, were 12-20 mm long and of area 39-111 mm^2^, equivalent to a circular diameter of ca 7-12 mm. Since typical clot fibril diameters are more like 100 nm (Pretorius et al. 2016) this implies further that they have been formed via the accretion of a great many fibres.

Other means of discriminating clot properties in terms of the outcomes following an ischaemic stroke include radiomics (Cahalane et al. 2021; Patel et al. 2023), transcriptomics (Santo et al. 2024) and proteomics (Desilles et al. 2022; Lopez-Pedrera et al. 2023); it will be of interest to see which elements of these may underlie the fibrinaloid phenotype. Penn *et al*. have sought proteomic biomarkers of stroke. Their first study of the clot proteome (Penn et al. 2018c) found proteins involved in inflammation (47%), coagulation (40%), and atrial fibrillation (7%), among others, atrial fibrillation being well associated with stroke risk (Kell et al. 2024), as well as a prothrombotic state (Bartus et al. 2020; Glowicki et al. 2019; Mołek et al. 2022). Their later studies (SpecTRA (Penn et al. 2018a; Penn et al. 2018b)) included thrombospondin and VWF as part of a biomarker panel. Validation studies showed that both thrombospondin and apolipoprotein B contributed; both had been seen as amyloidogenic markers in fibrinaloid microclots (Kell and Pretorius 2024), with thrombospondin – despite its modest concentration in both plasma (Buda et al. 2019; Kalaidopoulou Nteak et al. 2024) and in normal clots (Ząbczyk et al. 2019) – having the potential to be incorporated into the fibrin strands themselves (Bale 1987; Bale and Mosher 1986a, b; Bale et al. 1985)

Staessens *et al*. (Staessens et al. 2021a; Staessens et al. 2021b) and others (Kitano et al. 2022) also found neutrophil extracellular traps (NETs) in the thrombus taken from ischaemic strokes, where they were related to the severity (Denorme et al. 2022); NETs are also an important feature of fibrinaloid microclots (Pretorius et al. 2024), and were found in prothrombotic clots in the studies of Ząbczyk *et al*. (Ząbczyk et al. 2020). In particular, lower levels of extracellular DNA were related to the ease of thrombectomy (Vandelanotte et al. 2024).

Note that we are here focused on standard ischaemic stroke, though we recognise the existence of cerebral amyloid angiopathy (Jäkel et al. 2022; Theodorou et al. 2023), which is a type of cerebrovascular disorder that is indeed known to be characterized by the accumulation of amyloid beta-peptide within cerebral blood vessels.

Although this was a small sample, it is clear, as is recognised by other methods (Boodt et al. 2021; Goebel et al. 2020; Huang et al. 2022; Huang and Bhaskar 2022; Jolugbo and Ariëns 2021; Staessens and De Meyer 2021; Zhang et al. 2024b), that the clots are themselves highly heterogeneous, both within and between samples.

The recanalization rates of intravenous thrombolytics range between 30-60% in the first 6-24 hours, though the reasons for such poor rates are not entirely known (Molina et al. 2001). Here the clots remained as amyloid in nature despite the application of recombinant tPA (‘alteplase’) in 6 of the 8 patients. Despite the effectiveness of thrombectomy, futile recanalization, in which patients achieve successful recanalization but fail to improve their functional outcome, is surprisingly common, and a variety of risk factors are known (Deng et al. 2023; Deng et al. 2022; Karimian-Jazi et al. 2024; Nie et al. 2018; Nogueira et al. 2024; Shen et al. 2023; Zhou et al. 2022). A biochemical marker that might be predictive of futile recanalization would be of significant value, and it is not unreasonable that fibrinaloid microclots might provide one, just as they have in sepsis (Schofield et al. 2024). Future studies will seek to address this.

## Acknowledgments

We thank Drs Nitika Rathi and Piyali Pal for basic analyses of the clots, Mr Khaja Syed for setting up the clot bank and facilitating the transfer of material to Liverpool University, Ms Sally Flintham for help with clot collection and transfer, Ms Helen Malone for assistance including the consenting, and Ms Marie O’Brien for skilled assistance with microtoming and dewaxing at the University of Liverpool.

## Supplementary Materials

None.

## Author Contributions

AC performed the thrombectomies. JMG performed the fluorescence microscopy. JHP contributed the statistical analysis of the images. DBK, EP, AS and MW contributed to the conceptualisation, analyses, funding acquisition, drafting, and final editing. All authors contributed to the drafting and final editing, and have read and agreed to the published version of the manuscript.

## Data Availability Statement

All relevant data are provided.

